# Apixaban following discharge in hospitalised adults with COVID-19: Preliminary results from a multicentre, open-label, randomised controlled platform clinical trial

**DOI:** 10.1101/2022.12.07.22283175

**Authors:** Mark R Toshner, Carrol Gamble, J Kenneth Baillie, Ashley Best, Emma Bedson, Judy Bradley, Melanie Calvert, Elin Haf Davies, Annemarie B Docherty, Efstathia Gkioni, Dyfrig A Hughes, Thomas Jaki, R Gisli Jenkins, Ashley Jones, Martin J Landray, Jonathan Mant, Daniel F McAuley, Peter JM Openshaw, Duncan Richards, Paul Wicks, HEAL-COVID Collaboration, Charlotte Summers

## Abstract

**Background:** The role of thromboprophylaxis in the post-acute phase of COVID-19 is uncertain due to conflicting results from randomised controlled trials and observational studies. We aimed to determine the effectiveness of post-hospital apixaban in reducing the rate of death and hospital readmission of hospitalised adults with COVID-19.

**Methods:** HEAL COVID is an adaptive randomised open label multicentre platform trial recruiting participants from National Health Service Hospitals in the United Kingdom. Here we report the preliminary results of apixaban comparison of HEAL-COVID. Participants with a hospital admission related to confirmed COVID-19 and an expected date of discharge in the subsequent five days were randomised to either apixaban 2.5 mg twice daily or standard care (no anticoagulation) for 14 days. The primary outcome was hospital free survival at 12 months obtained through routine data sources. The trial was prospectively registered with ISRCTN (15851697) and Clincialtrials.gov (NCT04801940).

**Findings:** Between 19 May 2021 and 21 November 2022, 402 participants from 109 sites were randomised to apixaban and 399 to standard care. Seven participants withdrew from the apixaban group and one from the standard care group. Analysis was undertaken on an intention-to-treat basis.

The apixaban arm was stopped on the recommendation of the oversight committees following an interim analysis due to no indication of benefit. Of the 402 participants randomised to apixaban, 117 experienced death or rehospitalisation during a median follow-up of 344·5 days (IQR 125 to 365), and 123 participants receiving standard care experienced death or rehospitalisation during a median follow-up of 349 days (IQR 124 to 365). There was no statistical difference in the rate of death and rehospitalisation (HR: 0·96 99%CI 0·69-1·34; p=0·75). Three participants in the apixaban arm experienced clinically significant bleeding during treatment.

**Interpretation:** Fourteen days of post-hospital anticoagulation with the direct oral anticoagulant apixaban did not reduce the rate of death or rehospitalisation of adults hospitalised with COVID-19. These data do not support the use of prophylactic post-hospital anticoagulation in adults with COVID-19.

**Funding:** HEAL-COVID is funded by the National Institute for Health and Care Research [NIHR133788] and the NIHR Cambridge Biomedical Research Centre [BRC-1215-20014*].

## Introduction

Whilst the acute effects of COVID-19 are now well described,^1,2^ evidence has emerged of serious longer-term complications occurring in the post-acute phase of the illness in a significant proportion of patients. COVID-19 results in a high incidence of cardiovascular and pulmonary complications that may have long-term consequences including venous thromboembolism,^3,4^ persistent inflammation,^5^ and pulmonary fibrosis.^6^ Despite extraordinary success in identifying therapies to treat the acute stage of the illness,^7-9^ there is little evidence to guide treatment in the post-acute phase resulting in wide variation in practice and use of treatments without evidence.

A retrospective cohort study of 47,780 individuals with acute COVID admitted to NHS hospitals in England prior to August 31, 2020, showed that 29·4% were readmitted and 12·3% died after discharge, within a mean follow-up period of 140 days;^10^ similar rates were reported in a separate study in the USA.^11^ The rate of hospital readmission in England was 3·5 times greater than that seen in a matched primary care cohort over the same time period (matched on demographic factors, comorbidities, BMI and smoking status).

Abnormalities in coagulation occur in severe and fatal COVID-19,^12,13^ and clotting factors and platelet activation are implicated in the pathogenesis of severe acute COVID-19.^14^ There is a high incidence of thromboembolic events during the post-acute phase of COVID-19.^3,4^ However, discrepancies in risk estimation and the potential harms of anticoagulant treatment result in considerable uncertainty in therapeutic decision-making.

In 2020, the National Institute for Health and Care Excellence undertook a rapid evidence review to address the question “what is the effectiveness and safety of pharmacological prophylaxis to reduce the risk of venous thromboembolism in adults who have received care for COVID-19?” They were unable to identify any research studies addressing this question but found 11 clinical guidelines that made clinical management recommendations in this area.^15^ Consequently, NICE recommended evidence was urgently required to inform this area of clinical practice. HEAL-COVID, a platform trial, sought to address this evidence gap by investigating the effect of post-hospital treatments in comparison with standard care on hospital-free survival in adults who had been initially hospitalised with COVID-19: The oral direct-acting anticoagulant apixaban was selected as one arm of the trial. Apixaban was selected based on its widespread availability, oral route of administration, well-described safety profile with limited need for dose adjustment and low risk of drug interactions.

## Methods

### Study design

HEAL COVID is an adaptive randomised open label multicentre multi-arm multi-stage platform trial. The study was designed using a platform structure that allows multiple different treatments to be evaluated simultaneously and new treatments can be added. Follow-up data were collected through data-linkage to routine clinical data sources, and by patient-entered data collected via an app/web-based system (ATOM5) or telephone calls from the central trial team.

Here, we report preliminary results of the routine clinical data sources of the apixaban comparison with standard care (other arms still in progress). Trial participants are recruited from NHS hospitals across England, Wales, Scotland, and Northern Ireland. Ethical approval for the trial was granted by the South Central – Berkshire Research Ethics Committee (21/HRA/0646). The trial protocol, statistical analysis plan and other trial documentation are publicly available at https://heal-covid.net/for-staff/staff-page/.

### Participants

Participants were eligible to be enrolled into HEAL-COVID if they were aged 18 years or over, were approaching the end of their hospital admission (estimated date of hospital discharge within five days) and had SARS-CoV-2 infection associated disease (positive SARS-CoV-2 test relating to this hospital admission). Written informed consent was obtained from participants or their legal representative.

Participants were excluded from enrolment in the overall platform if it was planned that they were to receive treatment for a COVID-19 related condition with one of the trial medications after hospital discharge, if they were not expected to survive 14 days after hospital discharge, or if they had a medical history that might, in the opinion of the attending clinician, put the participant at significant risk if they were to participate in the trial.

Additionally, participants were excluded from the apixaban comparison if they had a known hypersensitivity to the medication, a previous medical history of a significant complication or allergy with the medication or medication drug class, active clinically significant bleeding, Childs Pugh C or worse chronic liver disease, known pregnancy or breast-feeding, coagulopathy, a known lesion or condition considered by the investigator to be a significant risk factor for major bleeding, or if they had received long-term prehospital administration of any other anticoagulant for a non-COVID-19 indication and anticoagulation was planned to be continued after hospital discharge.

### Randomisation and masking

Randomisation occurred via an online system and used equal probability between all active treatments a given participant was eligible for, and standard care as the control arm (i.e., a patient that is eligible for two active interventions and control will be randomized 1:1:1 between the three arms). Random permuted block randomisation was generated using STATA version 14.

Patient allocations were irrevocably generated following confirmation of eligibility and completion of the central randomisation online form by a member of the site trial research team. Allocation concealment was ensured as the online system did not release the randomisation code until the participant had been recruited into the trial (after confirmation of eligibility had been completed). Following randomisation, the system provided the allocation and participant trial identifier, sending automated emails of the same for central monitoring purposes.

### Procedures

Participants were randomised to receive apixaban 2·5 mg twice daily for 14 days, or the usual standard care (no post-hospital anticoagulation) and followed up for a maximum duration of 12 months. The dose and duration of apixaban was selected by an independent group of experts (UK-COVID Therapeutic Advisory Panel) on the basis that post-hospital thromboprophylaxis would need to balance the risks and benefits of the intervention, and therefore wanted to minimise duration of treatment. The risk of thromboembolism is elevated for extended periods after COVID (and other viral respiratory infections) but is greatest in the 30 days after the onset of infection (16). Given that there is usually a delay of several days from onset of symptoms to hospital admission, and the median length of hospital stay accounts for a further 5-8 days, 14 days of post-hospital anticoagulation was therefore considered to cover the period of highest risk.

### Outcomes

The primary outcome measure of the HEAL-COVID platform trial is hospital free survival, collected via routine health data obtained from NHS Digital, eDRIS (Public Health Scotland), SAIL (Health and Care Research Wales) and the Northern Ireland Clinical Trials Unit. Use of routine clinical data, such as that provided by NHS Digital, has been shown to be suitable for use in clinical trials.^17^

Secondary outcomes include days alive and out of hospital within 60 days of randomisation, all-cause mortality, and hospital readmission after index hospital admission. Suspected Serious Adverse Reactions (SSARS) are recorded.

### Statistical analysis

Using a two-sided log-rank test with a significance level of 1% and a hazard ratio of 1·5, 362 events (re-hospitalisation or death) would need to be observed to achieve 90% power for a 1:1 allocation ratio comparing a single active treatment with standard care. Assuming no dropouts and an event rate of 25% in the standard care arm,^10^ a total of 1754 patients (877 per arm) would need to be recruited for one comparison. The significance level was adjusted to allow for multiplicity.

All comparative analyses were based on contemporaneously enrolled participants that were eligible to receive the treatment in the comparison. Analyses of accumulating data were performed at regular intervals for review by the Independent Data and Safety Monitoring Committee (IDSMC). The IDSMC were asked to advise whether the accumulated data from the trial, together with results from other relevant trials, justified continuing recruitment of further patients, or further follow-up.

A pre-specified interim analysis occurred when 181 events (i.e., 50% of the planned total) were observed across both arms of a comparison. In the case of a treatment effect at interim analysis indicating that the active treatment was no better than standard care, the IDSMC could recommend that recruitment to this treatment should stop. No early stopping for benefit was planned. The IDSMC could, at any point, determine there was a safety concern and require a treatment arm to be stopped immediately.

Statistical analysis was undertaken using SAS version 9.4, and the statistical analysis plan is available via https://heal-covid.net/for-staff/staff-page/ and is included in the supplementary material. Analysis was by the intention to treat principle; primary outcome results are presented as Hazard Ratio (HR) and 99% confidence intervals (99%CI). The p-value of the log-rank test is additionally presented. Censoring within the time to event analyses was applied to participants who did not experience the event of interest after 12 months of follow up or earlier where follow up is ongoing or participants withdraw. Secondary outcomes (all-cause mortality and hospital readmission after index hospital admission) are analysed as per the primary outcome. For the secondary outcome, the number of days alive and out of hospital within 60 days (for purposes of interim analysis) analysis is restricted to participants with 60 days follow up, and the median and interquartile range is presented with p-value obtained from Mann-Whitney U test.

It should be noted that there is a delay (which varies by devolved nation) in the details of hospital admissions and deaths becoming available. This means that whilst participants may have completed follow-up, additional events may be notified sometime later.

HEAL-COVID is registered with clinicaltrials.gov (NCT04801940) and ISRCTN (15851697).

## Results

Between 19 May 2021 and 21 November 2022, a total of 1194 participants were enrolled into the HEAL-COVID platform trial across 109 sites, of whom 801 were recruited to the apixaban comparison (n=402 apixaban, n=399 standard care; Figure 1).

**Figure 1:**
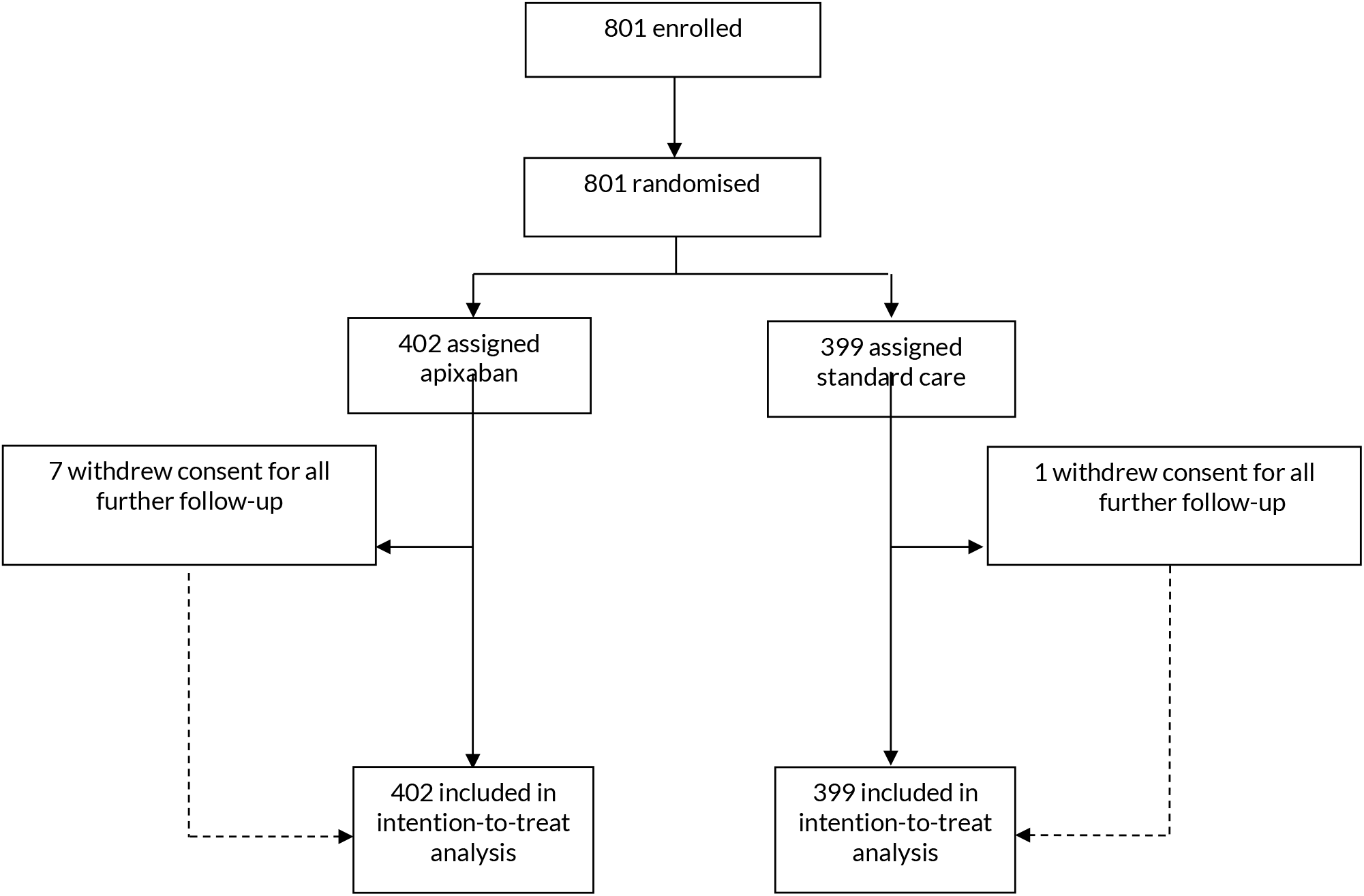
Trial profile.

The Independent Data Safety and Monitoring Committee met on three occasions, with the third meeting occurring when the pre-specified criteria to conduct the interim analysis were met for the apixaban comparison. The interim analysis used data obtained up to 20 October 2022. At this point, there were 797 participants (n=400 apixaban, n=397 standard care) and a total of 220 events (109 in apixaban and 111 in standard of care). On 7 November 2022, the IDSMC recommended to the Trial Steering Committee that the apixaban arm of the trial be discontinued due to there being no indication of benefit from apixaban for participants within the study. The Trial Steering Committee subsequently reviewed the interim analysis and on 19 November 2022 notified the Chief Investigator that the apixaban comparison should be stopped. The apixaban arm was closed to further recruitment on 21 November 2022, and an updated dataset of 801 participants analysed for the purposes of this manuscript.

The baseline characteristics of the participants are shown in Table 1. Of note, the mean age of the participants was 55·3 years, 59·8% had received one or more COVID vaccinations at the point they were enrolled into the trial, 61·9% were functionally unlimited prior to hospital admission, and 41·7% reported no comorbidities. Most participants (84·6%) required supplemental oxygen or other respiratory support and received drug therapy for acute COVID prior to being enrolled (84·4% dexamethasone/other corticosteroid, 25·3% remdesivir, 31·7% interleukin-6 receptor blockade).

**Table 1:**
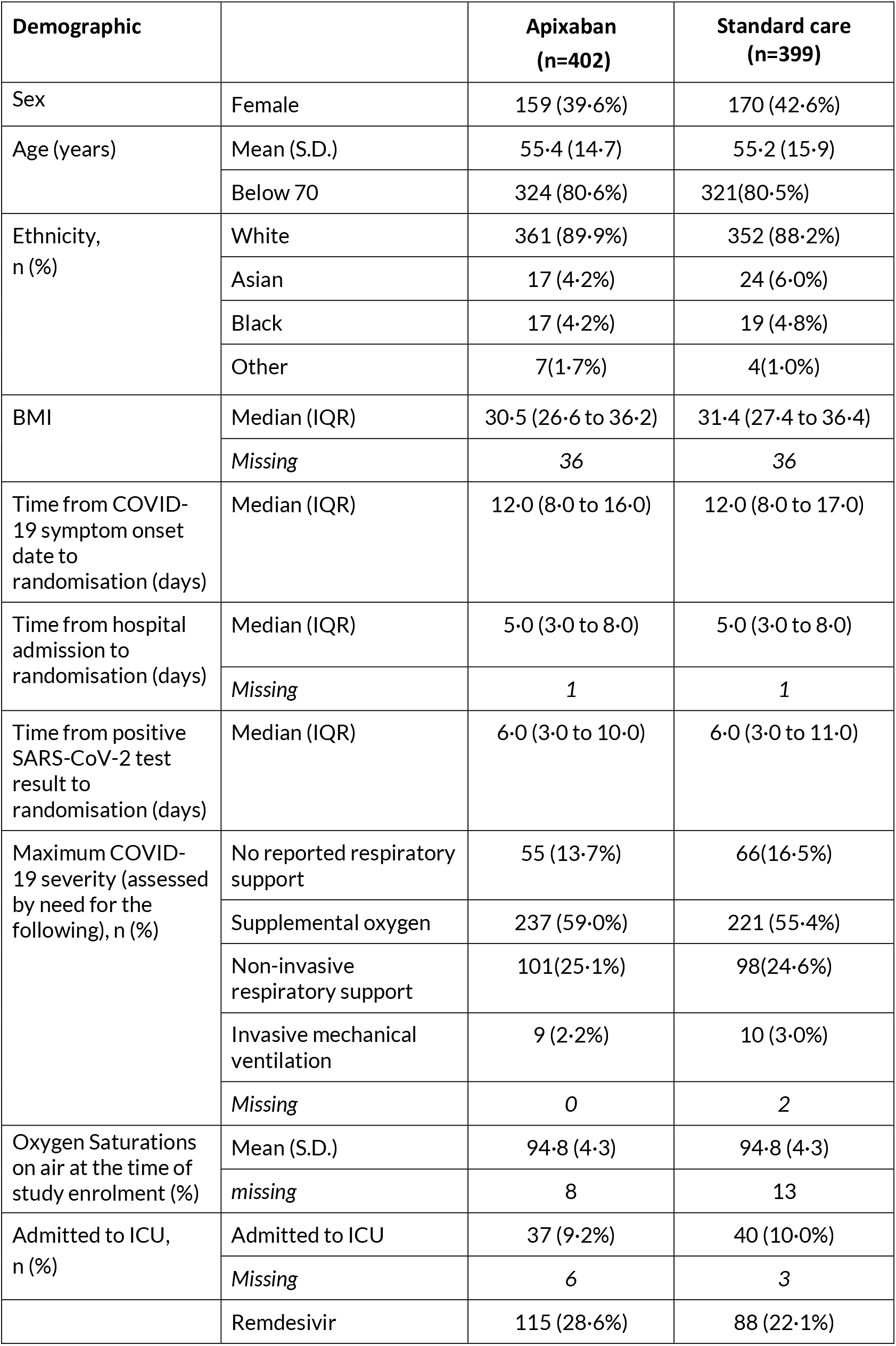
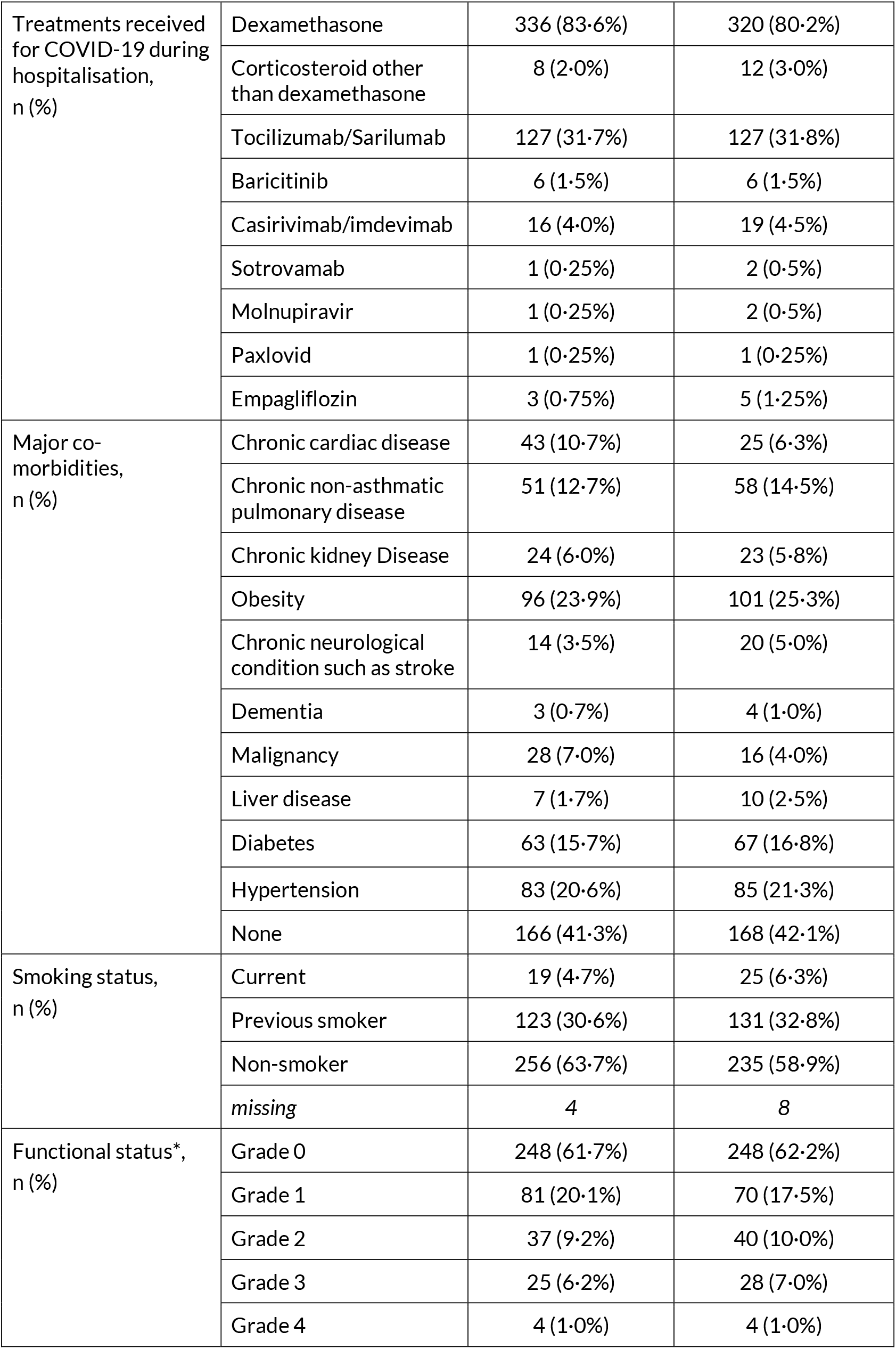
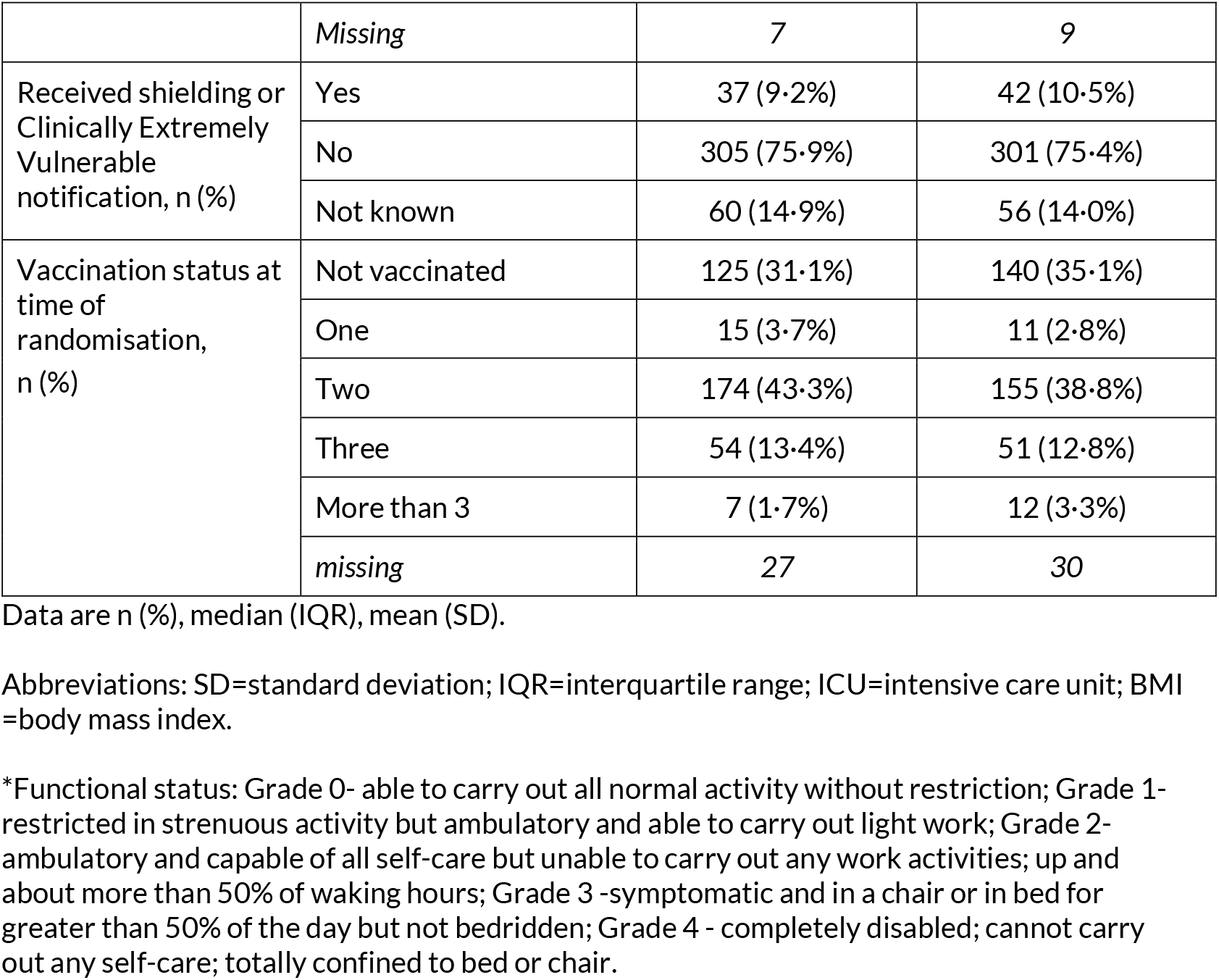
Baseline characteristics by treatment allocation.

There were 117 primary outcome events (death or readmission to hospital) in the apixaban group, compared with 123 events in those receiving standard care (HR 0·96, 99% CI 0·69-1·34), with a median follow-up period of 344·5 days (IQR: 125 to 365 days) and 349 days (IQR: 124 to 365 days) respectively (see Figure 2 and Table 2). Most of the events were readmissions (see Table 2). Twelve participants who were readmitted to hospital subsequently died during their hospital stay, so for the purposes of the primary endpoint were recorded as hospital readmissions.

**Figure 2:**
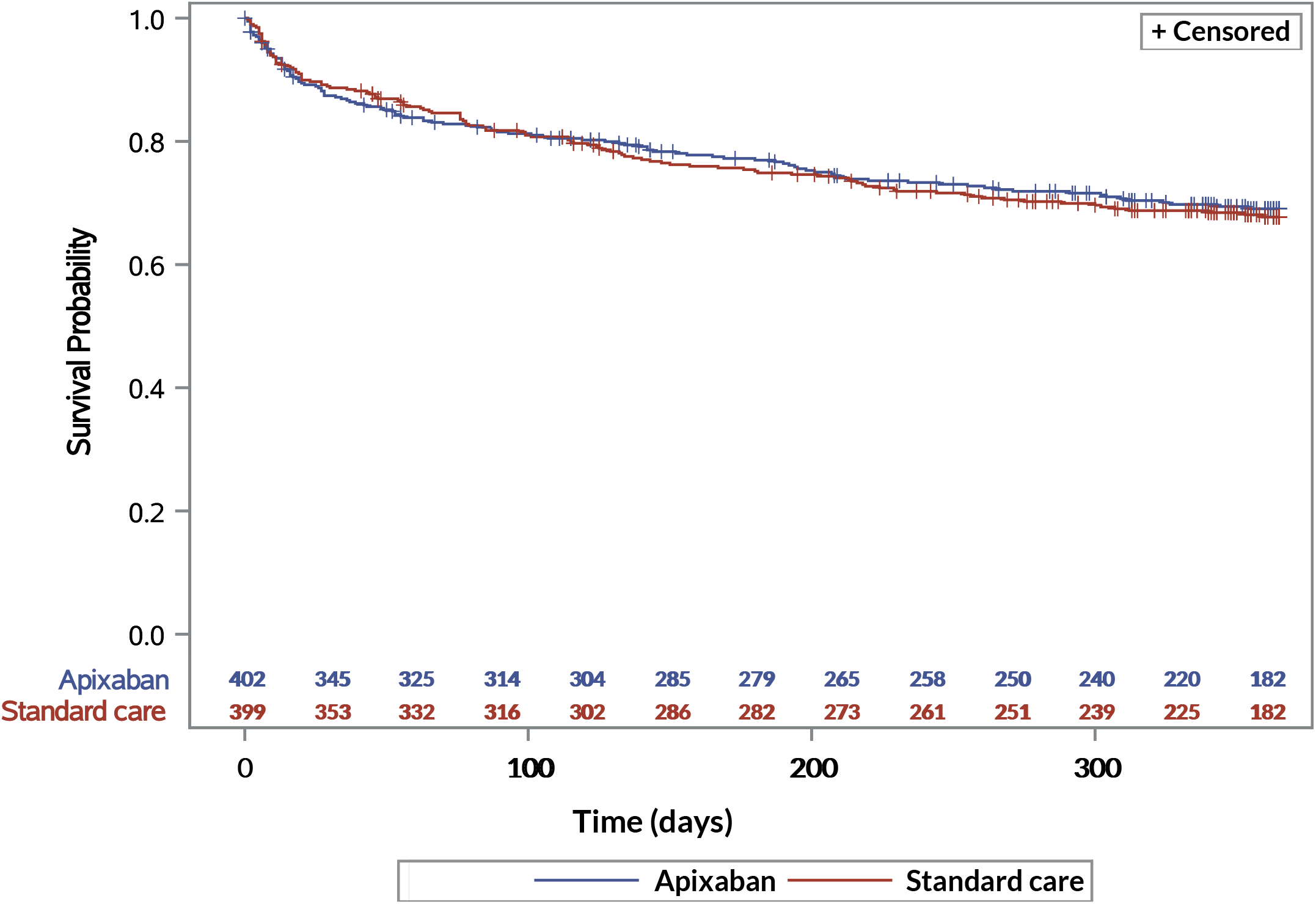
Effect of randomisation to apixaban on hospital-free survival.

**Table 2:** Effect of treatment allocation on main study outcomes.

|  | Apixaban<br>(n=402) | Standard care<br>(n=399) | HR<br>(99% CI) | Test<br>*p*-value |
| --- | --- | --- | --- | --- |
| Primary Outcome |  |  |  |  |
| Hospital free survival | 117 | 123 | 0.96 (0.69, 1.34) | Log-rank<br>*p*=0.75 |
| Death | 4 | 6 |  |  |
| Readmission | 113 | 117 |  |  |
| Secondary outcomes |  |  |  |  |
| Median (IQR) days alive and out of hospital 60 days after randomisation* | 59 (57-60),<br>n=385 | 59 (57-60)<br>n=388 |  | Mann<br>Whitney<br>*p*=0.66 |
| All-cause mortality<br>** | 10 | 12 | 0.83 (0.28, 2.51) | Log-rank<br>*p*=0.67 |
| Readmission to hospital | 113 | 117 | 0.98 (0.69, 1.37) | Log-rank<br>*p*=0.85 |

Data are n (%), median (IQR), mean (SD) or n/N (%).
Abbreviations: SD=standard deviation; IQR=interquartile range; SSARS= Suspected Serious Adverse Reactions' NA= Not applicable
\* Includes participants with minimum of 60 days follow up.
\*\* 12 patients who were readmitted to hospital subsequently went on to die, but only the first event contributes to the primary outcome.

There was no difference in the number of days alive and out of hospital within 60 days after randomisation between the apixaban and standard care group (median 59 days, IQR 57-60 versus 59 days IQR 57-60; p=0·66). There were 22 deaths: 10 in the apixaban arm compared with 12 in those receiving standard care HR 0·83 99% CI (0·28, 2·51).

Two participants who received apixaban experienced bleeding requiring hospital-based intervention and early cessation of the treatment (reported as SSARs). A third participant, identified within the routine data source, required hospital admission due to haemoptysis on day 11 after randomisation into the apixaban arm. The reported primary causes for death and hospital readmission during the 12 months after randomisation are shown in Tables 2 and 3 respectively. Of note, the four pulmonary emboli occurring in the standard care group all arose within 21 days of hospital discharge (Supplemental Table 1).

**Table 3:** Causes of death by treatment allocation.

|  | **Apixaban** | **Standard care** |
| --- | --- | --- |
| **Causes of death:** |  |  |
| COVID-19 | **3** | **6** |
| Cardiovascular | **2**<br>*Phlebitis and thrombophlebitis of other deep vessels of lower extremities (n=1)*<br>*Atherosclerotic heart disease (n=1)* | **1**<br>*Stroke, not specified as haemorrhage or infarction (n=1)* |
| Genitourinary | **2**<br>*Acute tubulo-interstitial nephritis (n=1)*<br>*Urinary tract infection, site not specified (n=1)* | **0** |
| Musculoskeletal | **0** | **1**<br>*Malignant neoplasm of connective and soft tissue (n=1)* |
| Respiratory (non-COVID) | **2**<br>*Chronic obstructive pulmonary disease with acute exacerbation, unspecified (n=1)*<br>*Chronic obstructive pulmonary disease, unspecified (n=1)* | **3**<br>*Chronic obstructive pulmonary disease with (acute) lower respiratory infection (n=1)*<br>*Chronic obstructive pulmonary disease, unspecified (n=1)*<br>*Type one respiratory failure due to pulmonary fibrosis (n=1)* |
| Neurological | **0** | **0** |
| Unknown | **1** | **0** |
| Missing | **0** | **1** |

Data are number of cases. Total cases for each group in bold, with specific individual causes listed in italics below.

**Table 4:**
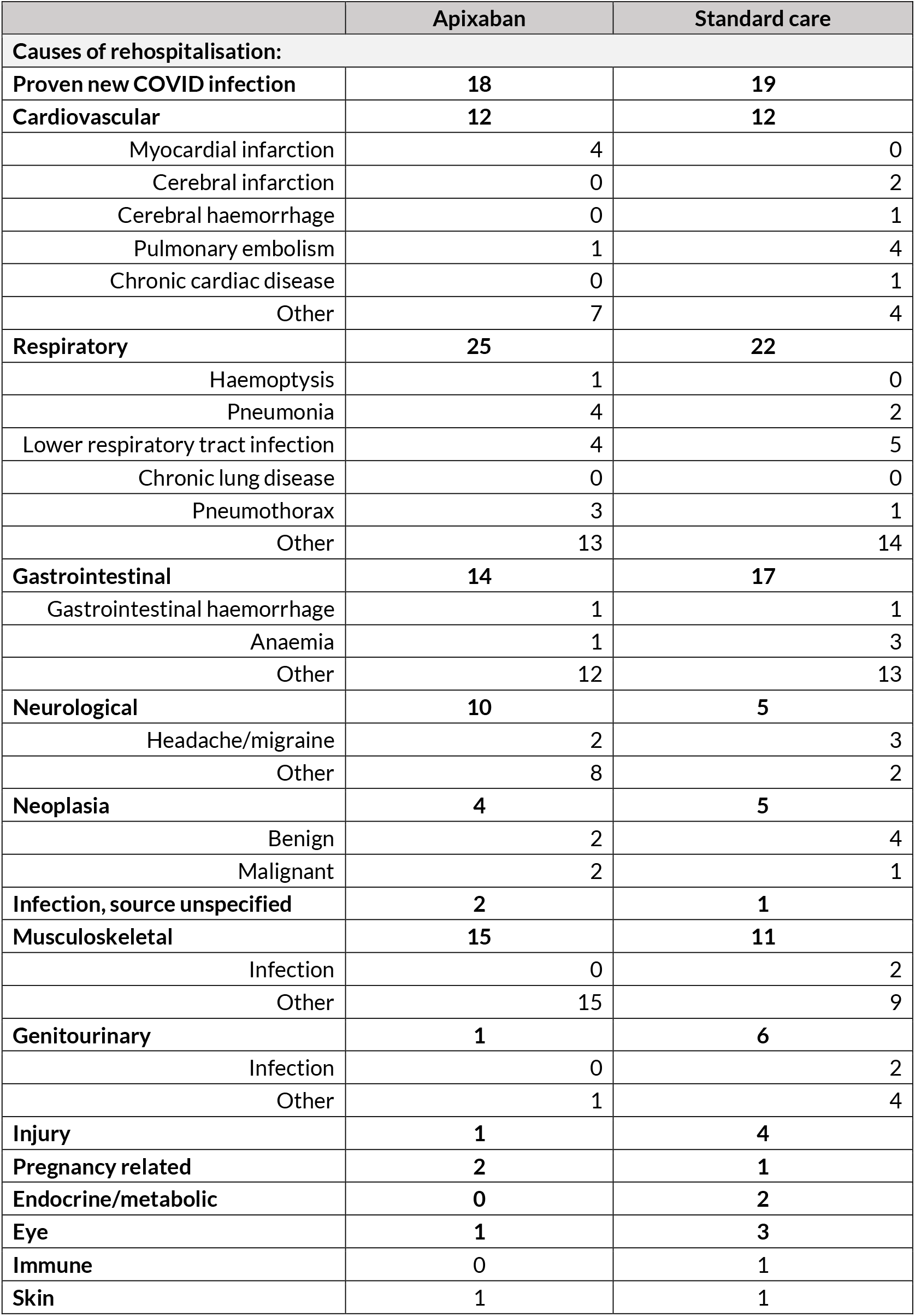
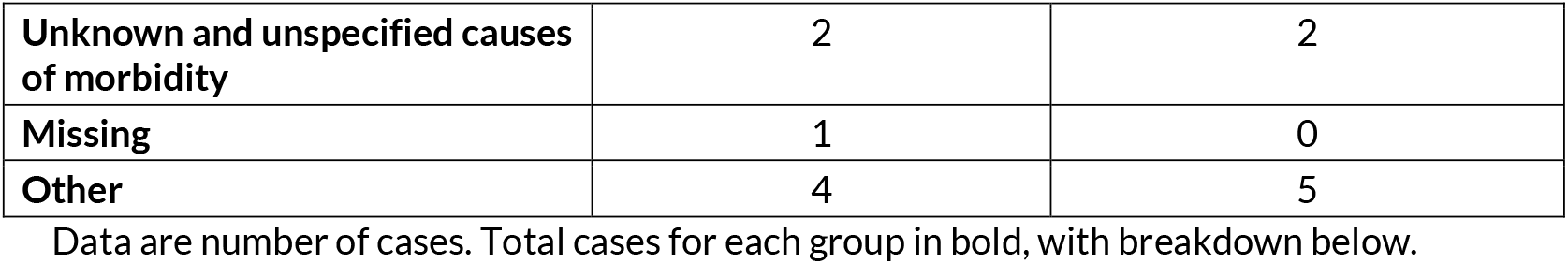
Causes of rehospitalisation by treatment allocation.

|  | Apixaban | Standard care |
| --- | --- | --- |
| **Causes of rehospitalisation:** |  |  |
| **Proven new COVID infection** | **18** | **19** |
| **Cardiovascular** | **12** | **12** |
| Myocardial infarction | 4 | 0 |
| Cerebral infarction | 0 | 2 |
| Cerebral haemorrhage | 0 | 1 |
| Pulmonary embolism | 1 | 4 |
| Chronic cardiac disease | 0 | 1 |
| Other | 7 | 4 |
| **Respiratory** | **25** | **22** |
| Haemoptysis | 1 | 0 |
| Pneumonia | 4 | 2 |
| Lower respiratory tract infection | 4 | 5 |
| Chronic lung disease | 0 | 0 |
| Pneumothorax | 3 | 1 |
| Other | 13 | 14 |
| **Gastrointestinal** | **14** | **17** |
| Gastrointestinal haemorrhage | 1 | 1 |
| Anaemia | 1 | 3 |
| Other | 12 | 13 |
| **Neurological** | **10** | **5** |
| Headache/migraine | 2 | 3 |
| Other | 8 | 2 |
| **Neoplasia** | **4** | **5** |
| Benign | 2 | 4 |
| Malignant | 2 | 1 |
| **Infection, source unspecified** | **2** | **1** |
| **Musculoskeletal** | **15** | **11** |
| Infection | 0 | 2 |
| Other | 15 | 9 |
| **Genitourinary** | **1** | **6** |
| Infection | 0 | 2 |
| Other | 1 | 4 |
| **Injury** | **1** | **4** |
| **Pregnancy related** | **2** | **1** |
| **Endocrine/metabolic** | **0** | **2** |
| **Eye** | **1** | **3** |
| **Immune** | **0** | **1** |
| **Skin** | **1** | **1** |
| **Unknown and unspecified causes of morbidity** | 2 | 2 |
| **Missing** | 1 | 0 |
| **Other** | 4 | 5 |

536 Data are number of cases. Total cases for each group in bold, with breakdown below.

We undertook *post hoc* analyses to determine whether there were differences in the primary outcome between several pre-specified subgroups and found no difference in the primary outcome with sex, age, body mass index, COVID severity or vaccination status at the point of randomisation (Figure 3).

**Figure 3:**
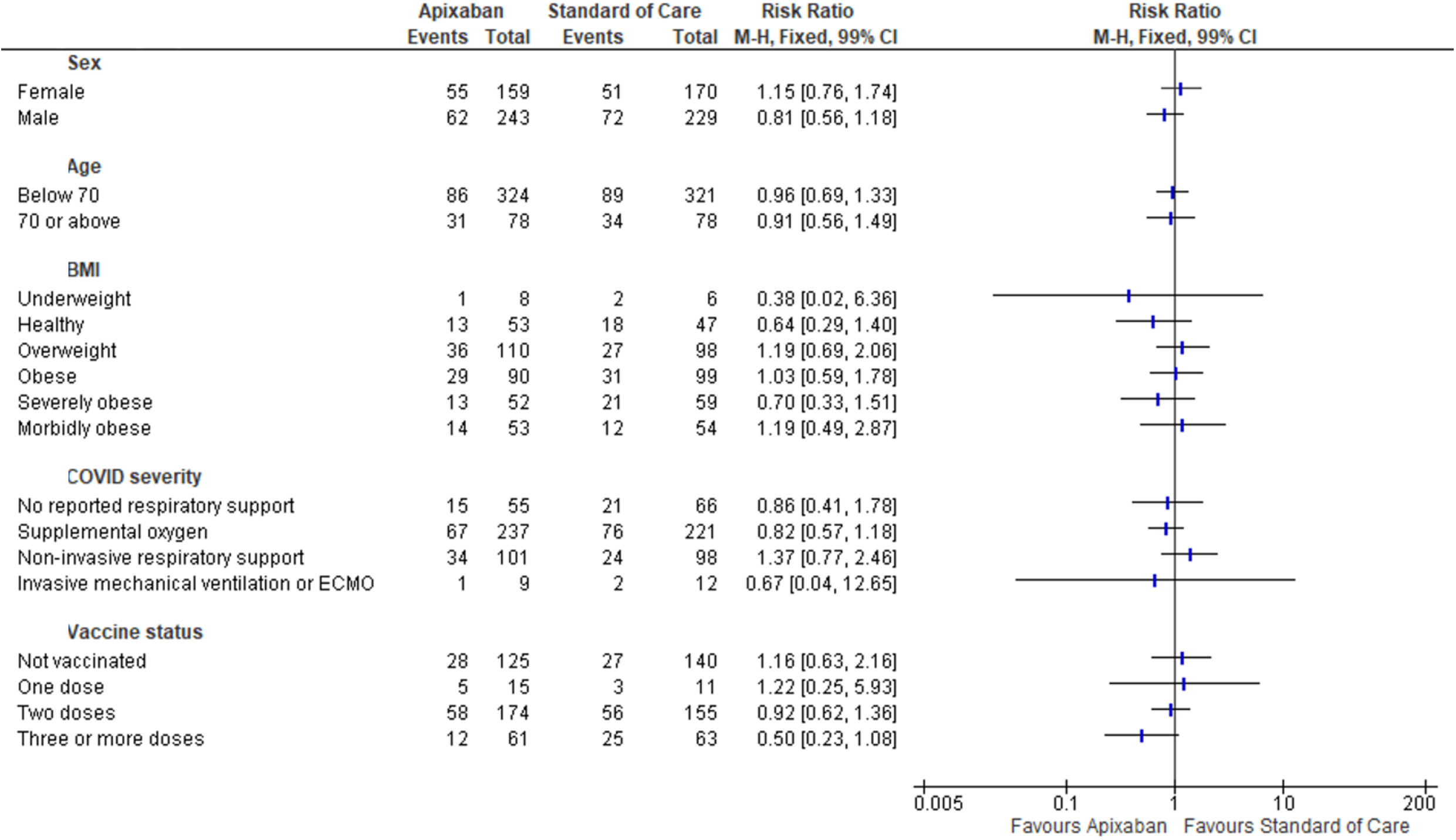
Effects of apixaban on event rate by baseline characteristics.

## Discussion

Our data show that treatment with 14 days of apixaban after hospital admission due to COVID-19 did not reduce the subsequent rates of death or hospital readmission. We have also shown that the rate of readmission to hospital remains unchanged from that seen prior to the introduction of vaccination and therapies such as dexamethasone, tocilizumab, and antivirals (standard care arm: mortality 3%; readmission 29.3%). Given nearly 1 million people with COVID have so far been admitted to UK hospitals alone, the risk that approximately one in three of them is subsequently being readmitted to hospital within the following year represents a pressing public health issue.

Real-world evidence suggests that the highest relative risk of thromboembolism occurs in the first four weeks after COVID infection.^15,18^ HEAL-COVID has to date followed patients out to a median of >300 days and consistent with previous reports, most significant venous thromboembolic events occurred within the first 30 days of the trial. HEAL-COVID is the first trial to investigate anticoagulation in a predominantly vaccinated population, at a time of stable acute therapeutic options such as dexamethasone, tocilizumab, and antivirals, and in time periods covering more recent variants. It is notable that VTE events, though following the same temporal profile, are significantly lower than earlier in the pandemic.^12^

Four pulmonary emboli occurred in the standard care group during the first 30 days after randomisation and none in the apixaban group, suggesting that 14 days of post-hospital anticoagulation may be able to prevent these events. However, two SSARS and a hospital admission all due to bleeding requiring clinical intervention also occurred during the first 30 days in the apixaban group, highlighting that the risk-benefit ratio of post-hospital anticoagulation is finely balanced. Given this, it would be challenging with such a low thrombosis event rate to power a trial to demonstrate acceptable risk/benefit and HEAL-COVID has demonstrated follow-up out to a year with no overall improvement in mortality or hospital readmission measures.

The choice of anticoagulant - apixaban was carefully selected. This agent consistently shows a better bleeding safety profile than other agents in meta-analyses.^19^ The paucity of significant venous thrombotic events occurring after the treatment window in the standard care group strongly suggests that our results are generalisable to other direct oral anticoagulant drugs. On this basis, longer treatment was unlikely to deliver additional benefit and many alternative agents are associated with a higher risk of bleeding.

There are two other randomised controlled trials relevant to post-hospital anticoagulation, the ACTION trial,^20^ and the MICHELLE trial.^21^ The ACTION trial investigated the effect of extended anticoagulation in 615 hospitalised subjects who had COVID symptoms for up to 14 days and an elevated D-dimer concentration.^20^ Participants were randomised to therapeutic or prophylactic anticoagulation, commenced in hospital. Therapeutic anticoagulation continued until day 30 (even if discharged) and prophylactic anticoagulation was standard prophylaxis with low molecular weight or unfractionated heparin and discontinued at hospital discharge.

In-hospital therapeutic anticoagulation followed by rivaroxaban up util day 30 did not improve a composite of time to death, duration of hospitalisation or duration of supplemental oxygen use over 30 days. The trial also found no reduction in deep vein thrombosis, pulmonary embolism, myocardial infarction (MI), mortality or hospital readmission over the 30-day follow-up period. ACTION reported an increased incidence of major and clinically relevant non-major bleeding in the extended anticoagulation arm (8% versus 2%; RR 3·64 (95% CI 1·61-8·27, p=0·0010).

In contrast, the smaller MICHELLE trial was an open label multicentre trial of post-hospital rivaroxaban versus no anticoagulation examining the effect of extended thromboprophylaxis on symptomatic and asymptomatic venous and arterial thromboembolism in a high-risk population of patients, more than 50% of whom had been admitted to an intensive care unit.^21^ Five of 159 subjects allocated to rivaroxaban and 15 of 159 subjects allocated to no anticoagulation were found to have a composite primary outcome of symptomatic or fatal venous thromboembolism, asymptomatic venous thromboembolism detected by bilateral lower limb venous Doppler ultrasounds and CT pulmonary angiogram, symptomatic arterial thromboembolism (MI, non-haemorrhagic stroke, and cardiovascular death at day 35). MICHELLE reported that no major bleeding occurred in either group, and clinically relevant non-major bleeding occurred in two participants in each arm of the trial.

There are substantial differences between the HEAL-COVID and MICHELLE trials. The HEAL-COVID apixaban comparison was more than twice the size (N=801) of the MICHELLE trial and recruited a general hospitalised population rather than a high-risk subset. The significant primary outcome reported in MICHELLE was not prospectively defined in the study protocol and has been suggested to be subject to a high risk of bias.^22^ In contrast to both HEAL-COVID and ACTION, MICHELLE reported no major bleeding in either arm of the trial, and no increase in clinically relevant non-major bleeding in the anticoagulation arm, suggesting that the small size of the trial may have limited its ability to determine the risk-benefit ratio of anticoagulation. Of note rivaroxaban has been shown to have a higher rate of bleeding than apixaban in multiple patient cohorts.^19,23^

HEAL-COVID is an open label clinical trial and therefore has a potential risk of bias. To minimise the risk of bias, the primary outcome of death and readmission was extracted from routine clinical data sources, blind to treatment allocations. Though HEAL-COVID recruits in over 100 centres in diverse settings across the UK, the population recruited to the trial was younger than in previously reported hospitalised COVID-19 cohorts and with less co-morbidity.^10^ Older co-morbid populations would be expected to have an increased risk of bleeding; therefore risk/benefit is an important concept and requires a high burden of proof.

In conclusion, we report that 14 days of post-hospital anticoagulation with the direct oral anticoagulant apixaban did not reduce the rate of death or rehospitalisation of adults hospitalised with COVID-19, and these data do not support the use of post-hospital anticoagulation in adults with COVID-19.

## Supporting information

Membership of the HEAL-COVID Collaboration

## Data Availability

Deidentified patient data will be available after the primary results have been published. Requests will be reviewed by the Trial Management Group and where at all possible access will be granted.

https://www.heal-covid.net

## Contributors

CS, MT, CG, TJ, ML, DFM, RGJ, JKB, EHD, MC, DR, AMD, DH, PW designed the clinical trial.

EB, CS, MRT, CG had responsibility for the management of the clinical trial.

CG, EG, AB, AJ analysed the trial data.

CS wrote the first draft of the manuscript, with input from MRT and CG.

All authors contributed to the concept for the platform trial, critically revised the manuscript, had access to the data, and were responsible for the decision to submit for publication.

EG, AB, AJ have directly accessed and verified the underlying data reported in this manuscript.

## Acknowledgements

HEAL-COVID is funded by the National Institute for Health and Care Research [NIHR133788] and the NIHR Cambridge Biomedical Research Centre [BRC-1215-20014*].

The views expressed are those of the authors and not necessarily those of the NHS, the NIHR, or the Department of Health and Social Care.

TJ is supported by a grant from UK Medical Research Council (MC_UU_00002/14).

