## Supplementary Table 1 for "Apixaban following discharge in hospitalised adults with COVID-19: Preliminary results from a multicentre, open-label, randomised controlled platform clinical trial"

**Supplemental Table 1 – Timing of thromboembolic and bleeding events that were a primary cause of hospital admission**

| **Cardiovascular** | **Apixaban** | **SoC** |
|---|---|---|
| Myocardial infarction | **4** | 0 |
| *Timing* | *day 53* | |
| | *day 78* | |
| | *day 101* | |
| | *day 142* | |
| Cerebral infarction | 0 | **2** |
| *Timing* | | *day 230* |
| | | *day 312* |
| Pulmonary embolism | **1** | **4** |
| *Timing* | | *day 9* |
| | | *day 11* |
| | | *day 17* |
| | *day 187* | *day 19* |
| Cerebral haemorrhage | **0** | **1** |
| *Timing* | | *day 27* |
| **Respiratory** | **Apixaban** | ***SoC*** |
| Haemoptysis | **1** | **0** |
| *Timing* | *day 11* | |
| **Gastrointestinal** | **Apixaban** | ***SoC*** |
| Gastrointestinal haemorrhage | **1** | **1** |
| *Timing* | *day 49* | *day 353* |
