## Supplementary material for "Apixaban following discharge in hospitalised adults with COVID-19: Preliminary results from a multicentre, open-label, randomised controlled platform clinical trial": Membership of the HEAL-COVID Collaboration

*Writing group*

*Trial Management Group*

Charlotte Summers (Chief Investigator); Mark R Toshner (Co-Chief Investigator); Carrol Gamble; Thomas Jaki; Martin Landray; Daniel F McAuley; R Gisli Jenkins; J Kenneth Baillie; Elin Haf Davies; Duncan Richards; Melanie Calvert; Paul Wicks; Annemarie Docherty; Dyfrig Hughes; Jonathan Mant; Peter Openshaw, Judy Bradley, Emma Bedson.

*Independent Data and Safety Monitoring Committee (IDSMC)*

Duncan Young (Chair); Theodore Jack Iwashyna; Hannah Durrington; Siobhan Creanor

*Trial Steering Committee*

James Chalmers (Chair); Wei-Shen Lim; Katherine Sleeman; John Hurst; Matthew Sydes; Frances Mair, Antonica Fletton, Stevina Southwell

*HEAL-COVID Central Coordinating Team*

CAMBRIDGE: Charlotte Summers; Mark Toshner; Joseph Newman (API scheme lead); Central telephone team - Sarah Hewitt, Sarah Berry and Sara Stearn; Carrie Bayliss (sponsor representative); Rachel Slade (finance); Nicola Forber (contracting); Denise Pinto (contracting)

LIVERPOOL CLINICAL TRIALS CENTRE: Statistical team - Carrol Gamble, Ashley Jones, Ashley Best, Efstathia Gkioni, Michaela Brown, Dannii Clayton, Rachel Cooper. Trial management team - Emma Bedson, Rachael Dagnall, Helen Hickey, Chloe Donohue, Simon Winn; Information System and Data Management Team - Sharon Kean, Jonathan Gibb, Anthony Shorrock, Linda Kane; Clare Jackson, Janet Harrison, Michelle Girvan.

*NIHR East of England Clinical Research Network*

Anne Priest; Harley Bircher; Sallyanne Hurford; Bonnie Jackson

*Individual Site Teams*

(Listed in descending order of the number of participants recruited at the site).

PI = Principal Investigator

API = NIHR accredited Associate Principal Investigator

**University Hospitals Bristol And Weston NHS Foundation Trust, Bristol Royal Infirmary** E Stratton (PI), C Blair (API), R Davies (API), L Morgan (API), C Stewart (API), M Thake (API), C Woodman (API), S Brooks, J Willis

**Mid Yorkshire Hospitals NHS Trust, Pinderfields General Hospital** M Thirumaran (PI), S Boot (Co-I), A Dwarakanath (Co-I), J Quinn (Co-I), C Eng (API), H Shankar Kumar (API), S Buckley, E Denis, A Major, A Metcalfe

**Cambridge University Hospitals NHS Foundation Trust, Addenbrooke's Hospital**
J Fuld (PI), H P Mok (Co-I), R Bousfield (API), S Coscione (API), E Y Lim (API), T Mamarelis (API), J Agato, J Domingo, E Kourampa, V Mendoza, C Pasquale II, E Robisco-Diaz, J Sanchez

**Liverpool University Hospitals NHS Foundation Trust, Royal Liverpool University Hospital** P Hine (PI), A Atomode (API)

**Cardiff & Vale University LHB, University Hospital of Wales** J Underwood (PI), T Evans, SA Frayling, C Oliver

**Northumbria Healthcare NHS Foundation Trust, North Tyneside General Hospital** A Aujayeb (PI), S Shakir (API), A Tomlinson (API), J Bell, A Harriman, H Mckie, M Panteli, T Smith, G Waddell

**Salisbury NHS Foundation Trust, Salisbury District Hospital** J Cullis (PI), B Eapen, H Morgan

**University Hospitals Bristol And Weston NHS Foundation Trust**, **Weston General Hospital** E Stratton (PI), R Duncan (API)

**Guy's and St Thomas's NHS Foundation Trust, Guy's Hospital and St Thomas's Hospital** M Ostermann (PI), N Lumlertgul (API), AK Balasubramanian, T Bawa, A Brown, K Burns, A Davies, P, DelosSantosDominguez, B George, A Packham, D Wood

**Southern Health and Social Care Trust, Craigavon Area Hospital** R Convery (PI)

**University Hospitals Birmingham NHS Foundation Trust, Heartlands Hospital** G I Walters (PI), M Abdelwahab, M Bellamy, T Bellamy, E Birkhamshaw, L Breslin, E Butler, P Ellis, C Huntley, P Juru, M Lacson, F Moore, H Morgan, D Papakonstantinou, C Reilly, M Sangombe, S Sapele, I Shaukat, Z Ullah, L Wood

**University Hospitals Birmingham NHS Foundation Trust, Queen Elizabeth Hospital Birmingham** D Parekh (PI), S Chaudhri (API), MBK Niazi (API), M Bates, NA Haider, C McGhee

**University Hospitals Plymouth NHS Trust, Derriford Hospital** JP Corcoran (PI), G Marsh (API), M Mwadeyi

**Sheffield Teaching Hospitals NHS Foundation Trust, Northern General Hospital and Royal Hallamshire Hospital** AAR Thompson (PI), M Plowright (API), F Ahmed, J Cole, K Harrington, M Ilyas, C Jarman, R Kirk, A Lye, J McNeill, S Megson, H Newell, T Newman, L Nwafor, L Smart, P Wade, S Walker, L Watson

**Betsi Cadwaladr University LHB, Ysbyty Glan Clwyd** D Menzies (PI)

**Northern Lincolnshire and Goole NHS Foundation Trust, Scunthorpe General Hospital** L Ali (PI)

**West Hertfordshire Hospitals NHS Trust, Watford General Hospital** R Vancheeswaran (PI), C Ellis

**NHS Fife, Victoria Hospital** P Liu (PI), S Finch (Co-I), S Fowler, S Pirie

**Somerset NHS Foundation Trust, Musgrove Park Hospital** I Dragusin (PI), M Vaida (Co-I)

**Barts Health NHS Trust, The Royal London Hospital** H Kunst (PI)

**Hull University Teaching Hospitals NHS Trust, Hull Royal Infirmary** N Easom (PI), K Drury (API)

**Calderdale and Huddersfield NHS Foundation Trust, Calderdale Royal Hospital** A Biswas (PI), B Callow (API)

**Belfast Health and Social Care Trust, Belfast City Hospital, Mater Hospital and Royal Victoria Hospital** D F McAuley (Co-PI), J M Bradley (Co-PI), J Stewart (Co-I, API), M Bailey, A Balakrishnan Nair, K Carey, D Dawson, SJ Hanna, M McFarland, S McGinnity, C McNeill, M McQuaid, S Moor, S Murphy, K Smyth, R Stone, D Tweed, A Usher-Rea, B Wells

**United Lincolnshire Hospitals NHS Trust, Lincoln County Hospital** A Aslam (PI), G Phalod, SL Shephardson, R Spencer, S Tavares

**Manchester University NHS Foundation Trust, North Manchester General Hospital** A Ustianowski (PI), T Scoones (Co-I), K Kuriakose (API), G Lindergard

**Royal Surrey County Hospital NHS Foundation Trust, Royal Surrey County Hospital** K McCullough (PI), H J Abu, S Donlon, C Piercy, S Stone, E Tarr

**Liverpool University Hospitals NHS Foundation Trust**, **University Hospital Aintree** PS Albert (PI), J Doherty (API), N Nicholas

**NHS Grampian, Aberdeen Royal Infirmary** J Cooper (PI), A Nicolson (API)

**NHS Lothian, Royal Infirmary of Edinburgh** AJ Gray (PI), J Caffekey (Co-I), JW Dear (Co-I), SO Lynch (Co-I), S Brito-Mutunayagam (API), C Blackstock, C Cheyne, J Grahamslaw, R O'Brien, A Williams

**Epsom and St Helier University Hospitals NHS Trust, St Helier Hospital** R Macfarlane (PI), V Taylor (API), N Blanco, G Blows, L Evans, A Mathew, M Rebolledo

**The Queen Elizabeth Hospital King's Lynn NHS Foundation Trust, The Queen Elizabeth Hospital** A Baral (PI)

**Royal United Hospitals Bath NHS Foundation Trust, Royal United Hospital** J Suntharalingam (PI), Z Maseko, K White

**University College London Hospitals NHS Foundation Trust, University College Hospital** M Marks (PI), S Logan (Co-I), A Andrews, H Sorrell

**Bedfordshire Hospitals NHS Foundation Trust, Bedford Hospital** T H Chapman (PI), D Bagmane (PI), R Fronda, W Makinde, M Penacerrada, J Sarella, L Ylquimiche

**Wye Valley NHS Trust, Hereford County Hospital** IA Du Rand (PI), S Anderson, E Andrews, J Annett, A Baker-Hedges, A BicknellJ Birch, S Cakir, E Collins, H Gashau, S Gayle, K Hammerton, L Moseley, J Porteous, P Ryan, S Turner, N Waheed,

**East and North Hertfordshire NHS Trust, Lister Hospital** A Wilkinson (PI), A Sinha (API)

**Shrewsbury and Telford Hospital NHS Trust, Royal Shrewsbury Hospital** N Capps (PI), R Heinink (Co-I), K Ibison, S Jose, A Lacy-Colson

**Bedfordshire Hospitals NHS Foundation Trust, Luton & Dunstable Hospital** P Shetty (PI)

**Bolton NHS Foundation Trust, Royal Bolton Hospital** R Ahmed (PI), S Latham, E Mckenna, R Mistry, K Rhead

**University Hospitals of Derby and Burton NHS Foundation Trust, Royal Derby Hospital** T Bewick (PI), F Al-Arrayed (API)

**North Tees and Hartlepool NHS Foundation Trust, University Hospital of North Tees** K Conroy (PI)

**Northern Lincolnshire and Goole NHS Foundation Trust, Diana, Princess of Wales Hospital** O Khan (PI)

**Wrightington, Wigan and Leigh NHS Foundation Trust, Royal Albert Edward Infirmary** I Aziz (PI)

**NHS Greater Glasgow and Clyde, Royal Alexandra Hospital** J Patrick (PI)

**Shrewsbury and Telford Hospital NHS Trust, The Princess Royal Hospital** N Capps (PI), N Ahmad (Co-I), H Moudgil (Co-I), S Jose, J Nixon

**Royal Papworth Hospital NHS Foundation Trust, Royal Papworth Hospital (Papworth Everard)** M Davies (PI), J Newman (Co-I), A Alsaaty (API), I Boubriak (API), F Jarvis (API), K Kulathevanayagam (API), P Wild (API), M Ben M'Barek, K Paques

**University Hospitals Birmingham NHS Foundation Trust, Good Hope Hospital** M Saim (PI), H Pervaiz (Co-I), AD Vellore (Co-I), K Arifeen, D Lenton, K Misra, H Willis

**North West Anglia NHS Foundation Trust, Hinchingbrooke Hospital** R Buttery (PI)

**United Lincolnshire Hospitals NHS Trust, Pilgrim Hospital** P Zafeiris (PI), K Netherton

**Epsom and St Helier University Hospitals NHS Trust, Epsom Hospital** R Macfarlane (PI), V Taylor (API), N Blanco, G Blows, L Evans, A Mathew, M Rebolledo

**Swansea Bay University LHB, Morriston Hospital** I Blyth (PI), L O'Connell (API), C Davies, S Richards, J Travers
**North Bristol NHS Trust, Southmead Hospital** E Moran (PI), J Tomlins (API)

**NHS Greater Glasgow and Clyde, Queen Elizabeth University Hospital** C Berry (PI), A Morrow (Co-I), K Fallon

**South Tees Hospitals NHS Foundation Trust, The James Cook University Hospital** DR Chadwick (PI), P Lambert

**NHS Lothian, Western General Hospital** O Koch (PI), M Ke (Co-I), S Ferguson, A Shepherd

**Blackpool Teaching Hospitals NHS Foundation Trust, Blackpool Victoria Hospital** D McGhee (PI)

**NHS Greater Glasgow and Clyde, Glasgow Royal Infirmary** HK Bayes (PI), E McGarry (API), S Doig, A Dougherty, A Munro

**James Paget University Hospitals NHS Foundation Trust, James Paget University Hospital** J Patrick (PI)

**NHS Tayside, Ninewells Hospital** M Spears (PI), A Shaw (Co-I and API), S Mohammed, R Solstice

**Nottingham University Hospitals NHS Trust, Nottingham City Hospital** S Ryder (PI)

**Lancashire Teaching Hospitals NHS Foundation Trust, Royal Preston Hospital** S Gudur (PI), W Yune (API), J Mills, S Sowden

**Salford Royal NHS Foundation Trust, Salford Royal Hospital** D Green (PI), H Merrill (Co-I and API)

**Aneurin Bevan University LHB, The Grange University Hospital** T Szakmany (PI), S Fairburn (Co-I), T James, A Waters

**County Durham and Darlington NHS Foundation Trust, University Hospital of North Durham** F Khalil (PI), T Pagadala (Co-I), K Lekhak (API), V Allinson, M Birt, A Kay, N Kingston, K Potts

**Yeovil District Hospital NHS Foundation Trust, Yeovil District Hospital** A Broadley (PI), S Board

**Nottingham University Hospitals NHS Trust, Queen's Medical Centre** S Ryder (PI)

**Leeds Teaching Hospitals NHS Trust, St James's University Hospital** K Holliday (PI), A Ali, A Ashworth, C Favager, S James, E Wade

**Imperial College Healthcare NHS Trust, St Mary's Hospital** O Kon (PI)

**Buckinghamshire Healthcare NHS Trust, Stoke Mandeville Hospital** M Shahidi (PI), K Manso, A Ngumo

**London North West University Healthcare NHS Trust, Ealing Hospital** P Papineni (PI)

**Oxford University Hospitals NHS Foundation Trust, John Radcliffe Hospital** B Angus (PI)

**Mid Cheshire Hospitals NHS Foundation Trust, Leighton Hospital** D Fullerton (PI), A Burton (API), C Gabriel

**Medway NHS Foundation Trust, Medway Maritime Hospital** L Vincent-Smith (PI)

**The Rotherham NHS Foundation Trust, Rotherham District General Hospital** A Hormis (PI), L Zeidan, V Maynard

**County Durham and Darlington NHS Foundation Trust, Bishop Auckland General Hospital** A Abraham (PI)

**Bradford Teaching Hospitals NHS Foundation Trust, Bradford Royal Infirmary** P Whitaker (PI)

**Hywel Dda University LHB, Bronglais General Hospital** M Hobrok (PI)

**Hull University Teaching Hospitals NHS Trust, Castle Hill Hospital** N Easom (PI), K Drury (API)

**Calderdale and Huddersfield NHS Foundation Trust, Huddersfield Royal Hospital** V Chau (PI)

**NHS Lanarkshire, University Hospital Monklands** M Patel (PI), M McFadden

**London North West University Healthcare NHS Trust, Northwick Park Hospital** AM Whittington (PI), MS Wu (API)

**University Hospitals of Derby and Burton NHS Foundation Trust, Queen's Hospital, Burton** U Nanda (PI)

**Royal Free London NHS Foundation Trust, Royal Free Hospital** S Mandal (PI), K Florman (API)

**NHS Lanarkshire, University Hospital Hairmyres** M Patel (PI), R Hamil, B Welsh

**NHS Lanarkshire, University Hospital** Wishaw M Patel (PI), A Smith (Co-I), S Clements, S Marshall

**Western Health and Social Care Trust, Altnagelvin Area Hospital** M Kelly (PI)

**Northern Health and Social Care Trust, Antrim Area Hospital** P Minnis (PI), M Drain (Co-I), R Doherty (API), J Gallagher

**Mid and South Essex NHS Foundation Trust, Broomfield Hospital** A Moore (PI)

**North Cumbria Integrated Care NHS Foundation Trust, Cumberland Infirmary** C Graham (PI), T Wilson

**County Durham and Darlington NHS Foundation Trust, Darlington Memorial Hospital** S Kamaruddin (PI)

**University Hospitals of Leicester NHS Trust, Glenfield Hospital** RA Evans (PI), M Bourne

**Royal Brompton & Harefield NHS Foundation Trust, Harefield Hospital** W D-C Man (PI)

**Barts Health NHS Trust, Newham University** Hospital H Kunst (PI)

**The Princess Alexandra Hospital NHS Trust, Princess Alexandra Hospital** P Russell (PI)

**Portsmouth Hospitals University National Health Service Trust, Queen Alexandra Hospital** K Tariq (PI)

**Royal Berkshire NHS Foundation Trust, Royal Berkshire Hospital** F K Kavvoura (PI), A Pankhania (API)

**Royal Cornwall Hospitals NHS Trust, Royal Cornwall Hospital (Treliske)** D Browne (PI)

**Swansea Bay University LHB, Singleton Hospital** I Blyth (PI)

**NHS Lothian, St John's Hospital** S Lynch (PI), J Carruthers, C Cheyne, A McGeough

**North Cumbria Integrated Care NHS Foundation Trust, The West Cumberland Hospital** C Graham (PI), R Harper

**South Eastern Health and Social Care Trust, Ulster Hospital** J Elder (PI), F McElwaine (Co-I), C McManus (Co-I), S Ni Shandair (Co-I), A Somerville (Co-I), S Hagan, S Kelly, R Mains

**Barts Health NHS Trust, Whipps Cross Hospital** H Kunst (PI)

**Manchester University NHS Foundation Trust, Wythenshawe Hospital** S Ramjug (PI)

**Betsi Cadwaladr University LHB, Ysbyty Gwynedd** C Subbe (PI)
